# The Business of Quality: Evaluating the Impact of Healthcare Quality Improvements on Staff Numbers and Patient Utilization in Sub-Saharan Africa

**DOI:** 10.1101/2023.02.03.23285375

**Authors:** Gloria P. Gómez-Pérez, Daniëlla Brals, Aafke E. de Graaff, Ibironke Dada, Bonifacia Benefo Agyei, Peter Risha, Elizabeth Bonareri, John T. Dekker, Tobias F. Rinke de Wit, Nicole Spieker, Wendy Janssens

## Abstract

Every year an estimated 5 to 8 million people die in low- and middle-income countries (LMICs) due to poor-quality care. Although quality improvements in healthcare facilities in LMICs are well-possible with tailored implementation plans, costs are often mentioned as a prohibiting factor. However, if quality improvements increase trust among patients, this might translate into increased visits and higher revenues for providers and enable them to further invest in quality. This paper assesses the potential business case of quality improvements in Sub-Saharan Africa (SSA). It focuses on both the public and private sector since the latter provides at least half of all health services in SSA. The analysis is based on a dataset including multiple assessments of quality and business performance indicators for almost 500 health facilities in Tanzania, Kenya, Ghana, Nigeria, and other SSA-countries. We studied the association between changes in quality assessment scores and subsequent changes in numbers of patient visits and staff as proxies for business growth. We found that quality improvements significantly improved business performance indicators, but only for those facilities that had already reached a certain level of quality to begin with. These findings suggest an S-shaped relationship between quality and business performance, leading to the existence of a ‘low-quality trap’. Substantial financial investments might be needed initially to support facilities at the bottom of the distribution in reaching a basic level of quality, after which further quality investments may start translating into increased revenues, enhancing business performance.

**Key messages:**

- Millions of people die every year in low- and middle-income countries (LMICs) due to low quality of care
- To reach Universal Health Coverage, drastic quality improvements are essential
- Achieving quality of care in LMICs is possible but challenging because it requires substantial financial investments, specialized skills and sufficient human resources
- We found there is a business case for quality investments in SSA, as quality improvements are associated with a significant increase in the number of patient visits and staff over time, indicative of greater revenue streams and financial capacity
- Targeted financing programmes together with technical assistance to healthcare facilities are critical to drive quality investments, especially for facilities at the beginning of their quality improvement journeys

## INTRODUCTION

Access to healthcare in LMICs has improved considerably during the last decades, making poor-quality care a bigger barrier to reducing mortality than insufficient access. It has been estimated that 60% of deaths from conditions amenable to healthcare are due to poor-quality, with 8 million people in LMICs dying per year from conditions that should be treatable by a basic healthcare system. Apart from affecting patient outcomes, poor-quality care leads to unnecessary social and economic losses, lack of patient trust, waste of resources, and catastrophic expenditures (Kruk *et al*. 2018a). Increasing access to care will be ineffective in improving health outcomes and universal health coverage (UHC), as long as quality of care is not simultaneously addressed. However, improving quality of care is difficult in LMICs due to limited funding and human resources, low patient trust and empowerment, infrastructure is faltering, and poor regulation when substandard care is delivered. Moreover, nearly 40% of health care facilities in LMICs lack access to clean water, and nearly 20% lack sanitation (WHO 2018a), further reducing healthcare quality.

These challenges are aggravated in Sub-Saharan Africa (SSA) which has 14% of the world’s population (2017) but carries the highest disease burden worldwide measured in disability-adjusted life-years (DALYs) (Roser & Ritchie 2017). With less than 1% of global health expenditure, only 3% of the world’s health workers (Anyangwe & Mtonga 2007), and difficulties to ensure the level of infrastructure require to provide good quality care, SSA cannot provide even the most basic health care to a large proportion of its people, let alone ensure the level of quality needed for better health outcomes. Per capita public expenditure on health in some countries of SSA, measured by purchasing power parity (PPP), was below USD 40 in 2018 (2018), compared to a WHO-recommended minimum level of USD 86, and a level of at least USD 200 per capita investment needed to achieve significant improvements in financial protection of the population for expenditures on health (Jowett M 2016). In addition, the World Health Organization (WHO) estimates quality of care in SSA is at only 63% of what is feasible given its health expenditure, with marked variations between countries from 25% to 94% (WHO 2018b).

Although an estimated 50% of healthcare in Africa is provided by the private sector reaching both high and low income groups, with figures up to 77% in some SSA countries (WHO 2018b), official development assistance (ODA) has bypassed the private healthcare sector for many years. The WHO has recently recognized the private sector as a key partner in achieving UHC in Africa (World Health Organization 2019). The need to involve the private sector in health care provision has been further underscored during the COVID-19 pandemic.

To improve quality of care, both financial resources and capacity building are required. Financial institutions can help with initial investments (e.g. through digital loans)(MCF 2021), but financial sustainability requires facilities to keep their business afloat by securing sufficient and continuous patient numbers to ensure sustained revenue streams. Patient numbers in turn crucially depend on the quality of care provided to keep patients’ experience positive. If patients do not sufficiently value the provided services at the requested prices, utilization will be low. In addition, low quality of care hampers the empanelment of clinics in insurance agencies, further undermining facilities’ ability to both secure a substantial number of clients as well as financial accessibility for patients. These processes complete and perpetuate a vicious circle of low demand and poor supply (Onno P. Schellekens 2007; Spieker 2020).

To date no quantitative evidence exists from SSA on whether improved quality of care in public and private clinics indeed leads to increased patient numbers, and hence increased revenues and better business performance. Such evidence would strengthen the business case of quality improvement in Africa. This paper is the first to investigate the association between improvements in quality and in business performance indicators for private small- and medium-healthcare providers in Africa. We analyse changes in quality assessments for 491 public and private healthcare facilities in 7 countries in SSA and relate those to changes in financial growth, as exemplified by increased patient numbers and increased numbers of health care staff as proxies for business performance.

## MATERIALS AND METHODS

### SafeCare methodology

Our study focuses on clinics that participated in the SafeCare quality improvement program. The latter encompasses a quality improvement and stepwise certification approach developed in 2009 by PharmAccess Foundation, the Joint Commission International (JCI), and the Council for Health Service Accreditation of Southern Africa (COHSASA) to provide innovative health care standards, and a quality improvement process broken into achievable, measurable steps to facilitate incremental improvement of quality (Johnson *et al*. 2016). SafeCare has been built using a comprehensive set of ISQua (International Society for Quality in Health Care) accredited clinical standards that allow the evaluation and rating of healthcare quality in small- and medium-size facilities in LMICs. Its quality assessment is organized along 13 service elements that constitute the various medical and non-medical aspects of health care delivery, comprising a total of 753 SafeCare criteria and summarized into an overall score (Johnson *et al*. 2016). After a baseline assessment, a tailored quality improvement plan (QIP) is developed for each facility to guide the process of improvement. The QIP is built upon the identification of priorities for each healthcare facility. SafeCare also provides technical assistance to help achieve the QIP is provided. This is tailored to the context of each healthcare facility based on its financial and human resource capacities, and the baseline quality level. Finally, based on the SafeCare scores, each facility receives a rating ranging from level 1 (very modest quality) to level 5 (continuous quality improvement systems in place). The rating allows benchmarking between facilities and help providers to track their progress. It also provides policy-makers, users and donors in the healthcare system with information for informed decision making and resource allocation.

In addition to the SafeCare program, PharmAccess Foundation offers healthcare facilities access to small-and medium-sized loans through the Medical Credit Fund (PharmAccess 2022) to finance quality improvement implementations, as well as business support training.

### Study design and sampling

We conducted a retrospective cohort study to investigate the association between the changes in quality of care measured by SafeCare standards and the business performance of healthcare facilities as represented by the number of patient visits and number of medical staff. The research population consisted of 3089 health care facilities from Tanzania, Ghana, Kenya, Nigeria, Namibia, Liberia, and Uganda participating in the SafeCare program since 2011 and with at least one SafeCare assessment. Facilities were included in the analysis if they had ≥ 2 measurements of performance data that were ≥ 18 months apart (baseline and follow-up measurements), and SafeCare assessments that were performed no more than 3 months before or after the baseline performance data measurement, and no more than 3 months before or 6 months after the follow-up performance data measurement (Figure 1). In addition, secondary hospitals and facilities with high SafeCare scores were excluded from the sample since the magnitude of potential score improvement was very small, hence not contributing to the linear regression models (Figure 1). 491 facilities located were included in this analysis as these facilities had at least two performance measurements that were ≥ 18 months apart with matching SafeCare scores as per inclusion criteria and were below the level of secondary hospital or did not have high SafeCare scores at baseline (Figure 1).

**Figure 1.**
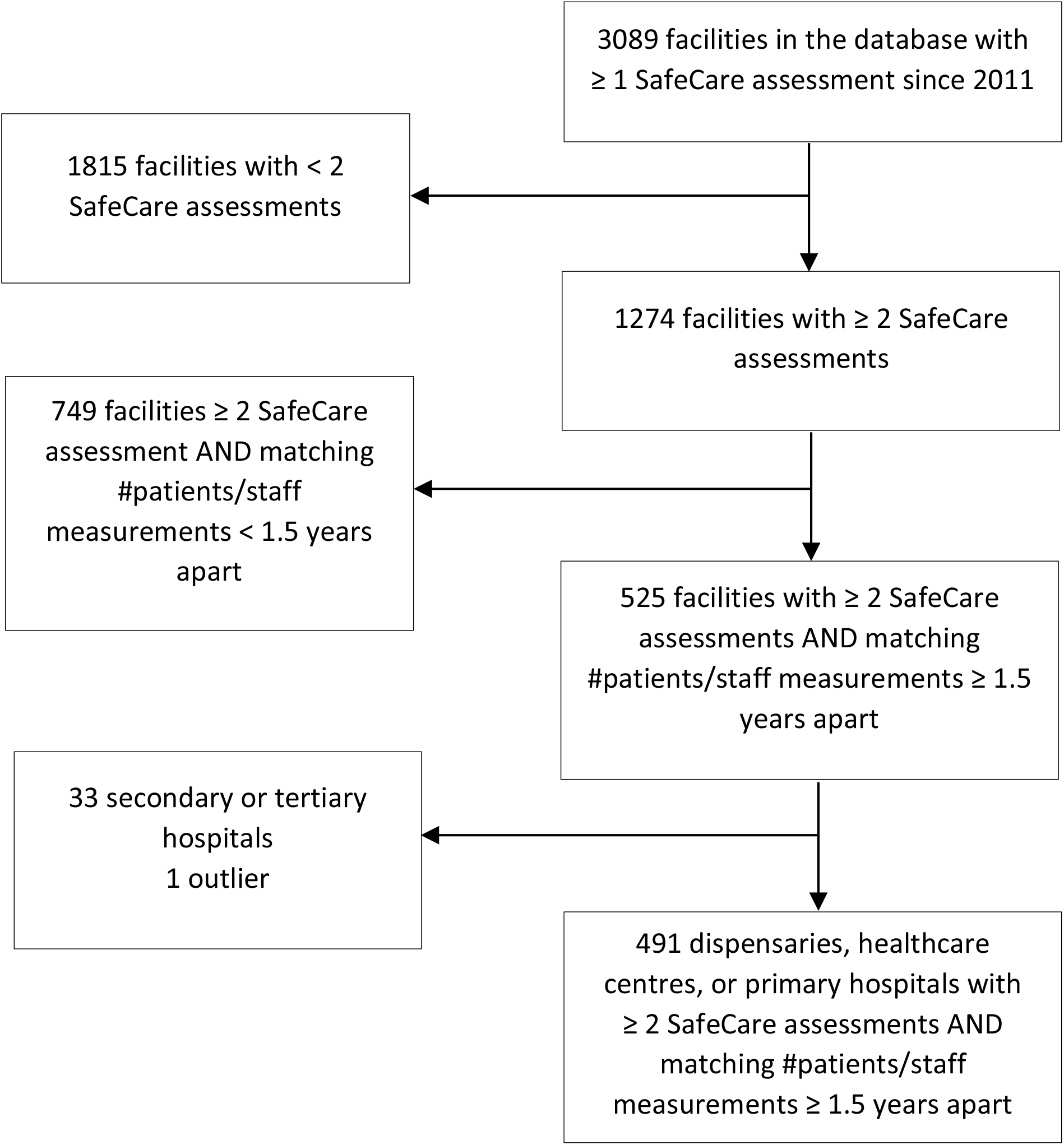
Study flowchart.

### Measurement of variables

The numbers of patient visits and medical staff, termed performance measures in this paper, consisted of the average number of monthly patients visits and medical staff reported by the facilities during the 6 months prior to collection date of performance measure data. Staff included nurses and medical officers, amongst others, with a part-time or full-time employment contract in the clinic at the time of the measurement. Quality improvements were measured as the change in SafeCare score between the baseline and follow-up measurements. After an assessment, a SafeCare score is calculated from 1-100. Scores represent a punctuation of all applicable criteria as fully compliant (2 points), partially compliant (1 point), or non-compliant (0,25 points) within each of the 13 service elements. More details have been published elsewhere (Johnson *et al*. 2016). Outcome variables were the change in the number of patients and the change in the number of staff between the baseline and follow-up measurements of performance data.

### Characteristics of facilities

491 facilities located in Tanzania, Kenya, Nigeria, Ghana, Uganda, Liberia, and Namibia were included in this analysis as these facilities had at least two performance measurements that were ≥ 1.5 years apart with matching SafeCare scores as per inclusion criteria and were below the level of secondary hospital at baseline (Figure 1).

### Statistical analysis

All analyses were performed using Stata version 16.1 (StataCorp). Facilities’ characteristics between baseline and follow-up were compared using the t-test for paired data. The quartiles of baseline quality score distribution were calculated to create four quality score categories as follows: low [15-36], lower-medium <36-44], higher-medium, <44-55], or high <55-94]. Associations between the change in SafeCare score and the change in the number of patients and staff were assessed in ordinary linear regression analyses, adjusting for the following (potential) confounders: quality score at baseline stratified by quality score categories (quartiles), number of patient visits/staff at baseline, facility level (dispensary, healthcare centre, or primary hospital), facility ownership (private, public, or faith-based), location (rural or urban), the number of days between the two SafeCare assessments that were used to calculate the change in score, and country that included five categories: Tanzania, Ghana, Kenya, Nigeria, and other. Data from Namibia, Liberia, and Uganda were merged under the category ‘other’ due to the small sample size of facilities included in the analysis per each of these countries. Same analysis was performed to study the association between the change in quality score only of the SafeCare criteria specifically related to business performance, comprising 38 out of 753 criteria, and the change in the number of patients and staff.

Next, we classified the facilities in improvers and non-improvers to compare the baseline characteristics and facilities’ profile. We performed heterogeneity analyses to calculate a quality improvement cut off from baseline to follow-up for this classification: improvers, ≥ 5 point-improvement from baseline; and non-improvers, < 5 point-improvement from baseline. Cut offs of 3-, 7-, and 10-point improvement were also tested showing the same associations than the 5-point cut off, but the latter was chosen because it showed the more significant differences between the two groups.

## RESULTS

### Description of the facilities

The analysis sample included 209 dispensaries (42.6%), 188 healthcare centres (38.3%), and 94 primary hospitals (19.1%). These were mostly of public ownership (n [%] 286 [58.2]), located in urban areas (313 [63.7]), and from Tanzania (256 [52.1]) and Kenya (169 [34.4]) (Table 1). At baseline, facilities received on average 1,119 patient visits per month and had 33 employees. These overall numbers increased to 1,227 visits per month at follow-up (at least 18 months later), but not significantly (*P* = 0.33). In addition, the average quality score among these facilities was within the higher-medium category at baseline (mean, 45.1) and improved significantly at the follow-up (mean, 56.1) (Table 1). Overall, we observed an improvement in the quality scores of participating facilities (Figure 2), and a larger proportion of facilities within the higher quality score categories (higher medium and high quartiles) at follow-up when compared to baseline (Table 1). Conversely, we observed that the proportion of facilities that at baseline were within the low and lower-medium quality score categories decreased significantly at the second time point (Table 1).

**Table 1.**
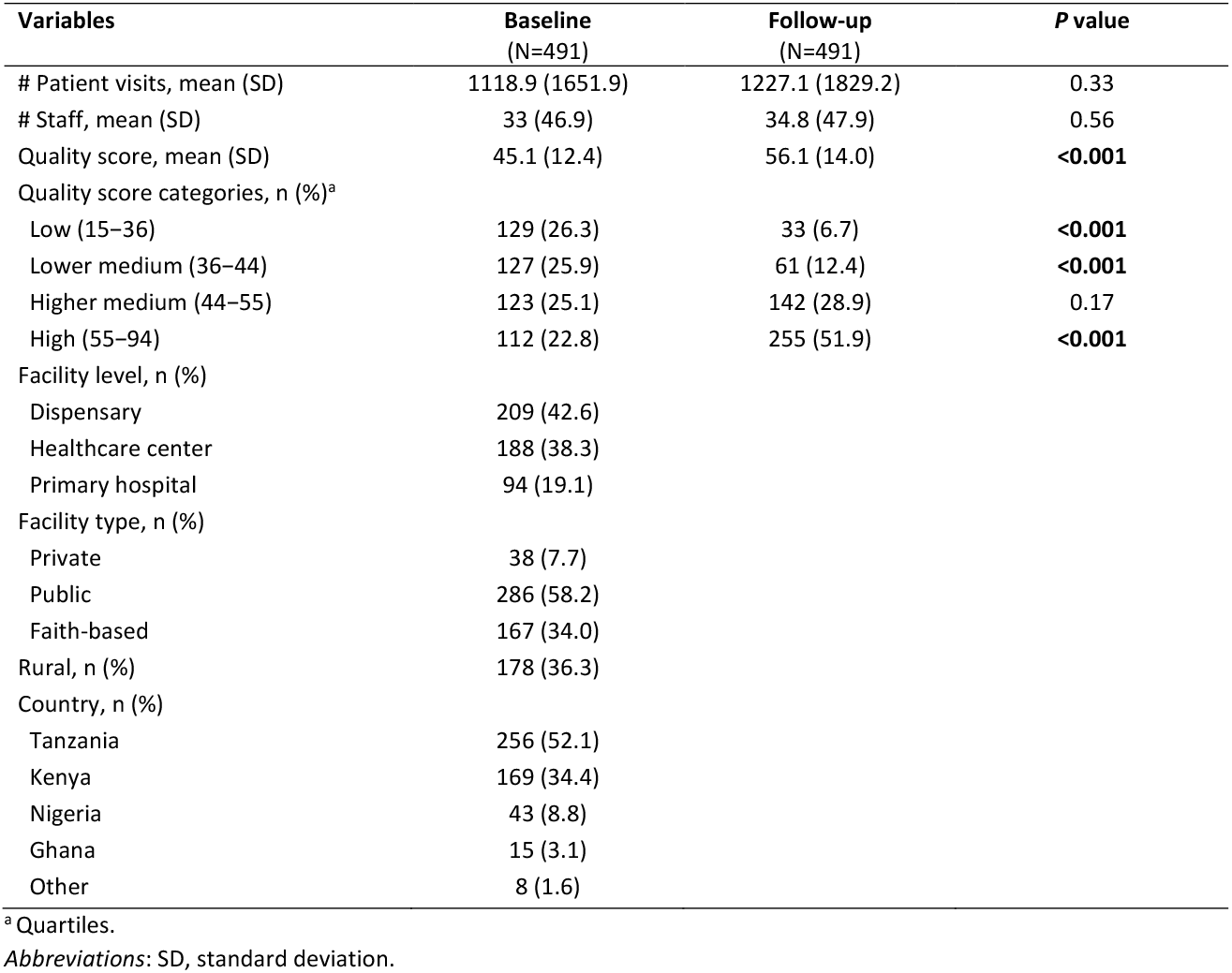
Facilities characteristics at baseline and follow-up.

**Figure 2.**
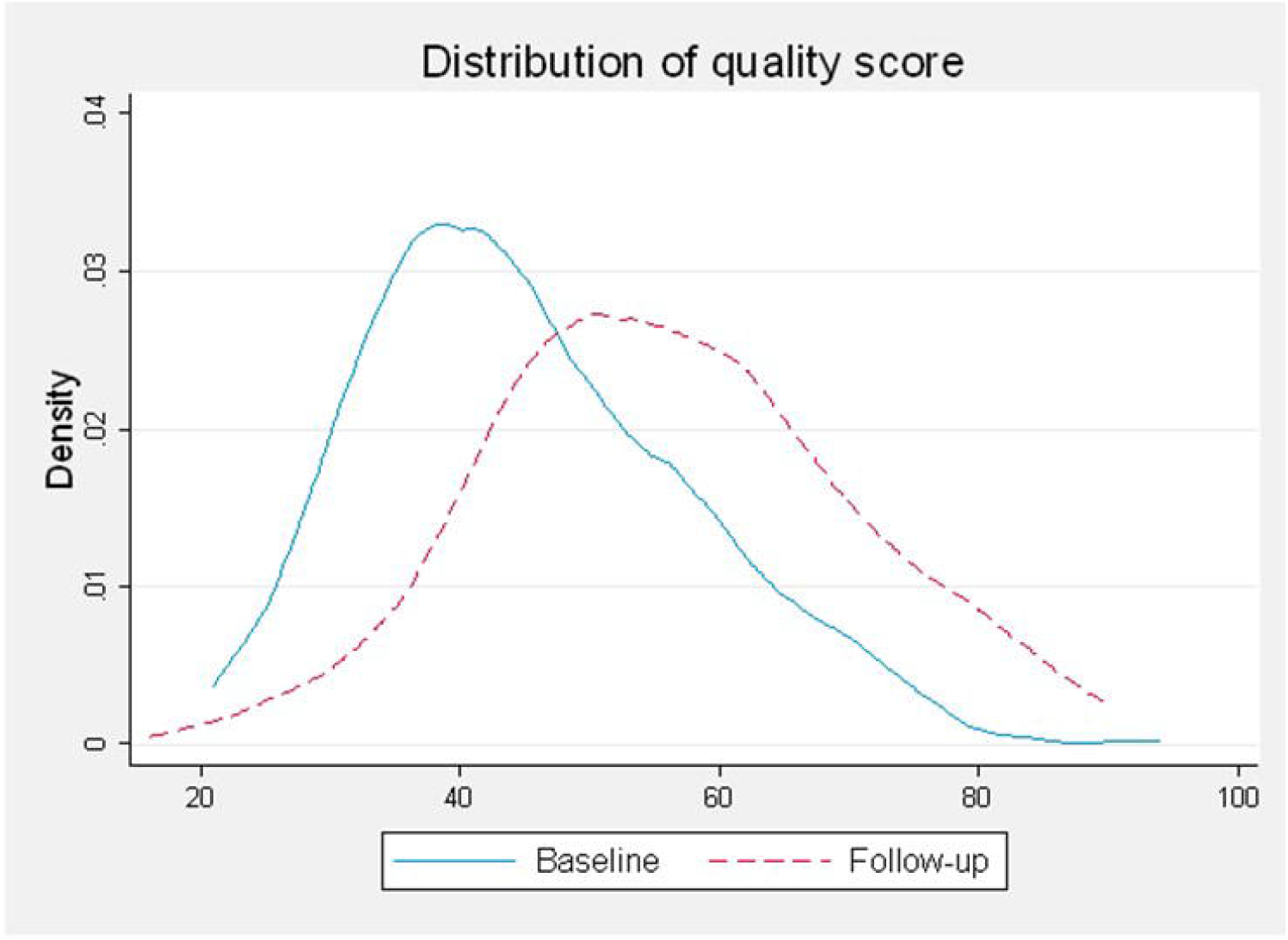
Distribution of quality score at baseline and follow-up. This figure shows that after ≥ 18 months follow-up, the proportion of facilities with a low and lower medium quality score (score <44) decreases, while the proportion of facilities with higher medium and high scores (score ≥ 44) increases, showing overall an improvement in quality in this group of facilities with time.

### Association between the change in quality score and change in the number of patient visits

We found a significant positive association between the change in quality score and the change in patient visits in these facilities (Table 2 Column ‘Ordinary linear regression’). On average, a 1-point increase in quality score was associated with an increase of 14.4 patient visits per month (95% CI 5.5–23.3, *P* = 0.002). This association remained significant after controlling for other possible predictor variables (Table 2 Column ‘Multiple linear regression’). According to this extended model, for each 1-point increase in SafeCare quality score, a facility could expect an average increase of 18 patient visits per month (95% CI 8.4-27.6, *P* < 0.001). Facilities that started with a high-quality score at baseline achieved significantly larger increases in the number of patients compared to facilities that started with a low-quality score (*P* = 0.04), irrespective of their subsequent change in quality score. Hospitals in comparison with dispensaries (*P* < 0.001), and urban facilities versus rural facilities (*P* = 0.04), showed significantly higher increases in patient numbers between baseline and follow-up. Additionally, the longer the follow-up time, the higher the observed increase in patient numbers. Heterogeneity analyses confirmed that only the facilities with a high quality baseline score saw a change in quality translated into an actual increase in the number of patient visits, and therefore into a better business performance (Table 2 Column ‘Heterogeneity analysis’). We also looked only at the score of the 38 business performance criteria and its association with change in patient visits. We found that the facilities that at baseline had a high-medium business performance score (assessment of 38 business performance criteria) were able to translate the improvements in business performance score into a higher number of patients visits (Supplementary Table 1).

**Table 2.**
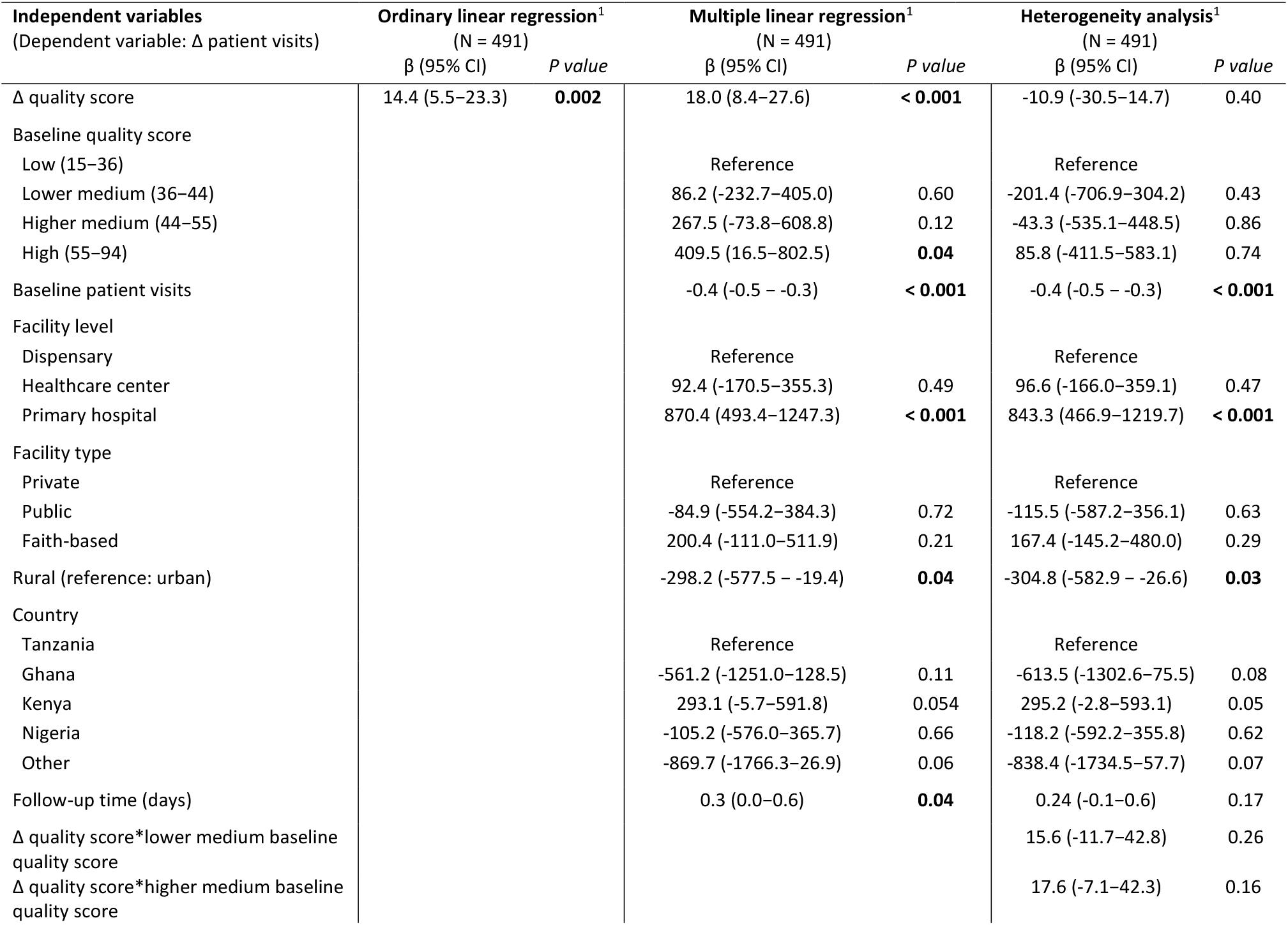
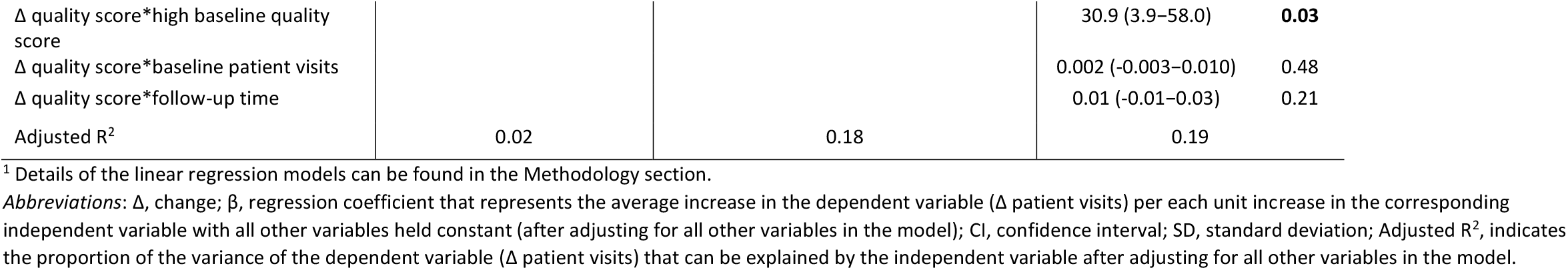
Linear regression models to estimate the effect of the change in quality score on the change in patient visits.

### Association between the improvement in quality score and the strengthening in the number of staff

Similar to the previous model, we also found a positive significant association between the improvement in quality scores and the increase in staff numbers (Table 3 Column ‘Ordinary linear regression’), that remained significant after controlling for other variables (Table 3 Column ‘Multiple linear regression’). For each 7-point increase in quality score a facility could achieve an average increase in 1 staff member (95% CI 0.28-1.82). Facilities with a high-quality score at baseline were better able to translate the improvement of quality into higher staff numbers compared to facilities with low baseline scores. Also, the growth in staff numbers associated with the improvement in quality care is most pronounced for hospitals. The change in staff numbers was not significantly associated to the ownership or the location of the facility, nor to the country or the time to follow-up. The heterogeneity analysis indicated that the larger facilities (higher staff numbers at baseline) were better able to translate quality improvements into changes in staff numbers (Table 3 Column ‘Heterogeneity analysis’). In addition, a trend was observed (*P* = 0.08) for the facilities that started with a high-quality score to translate quality improvements into an actual increase in the number of staff. Regarding the association between improvement in quality score of the 38 business performance criteria and the strengthening of staff numbers, we found similar results (Supplementary Table 2).

**Table 3.**
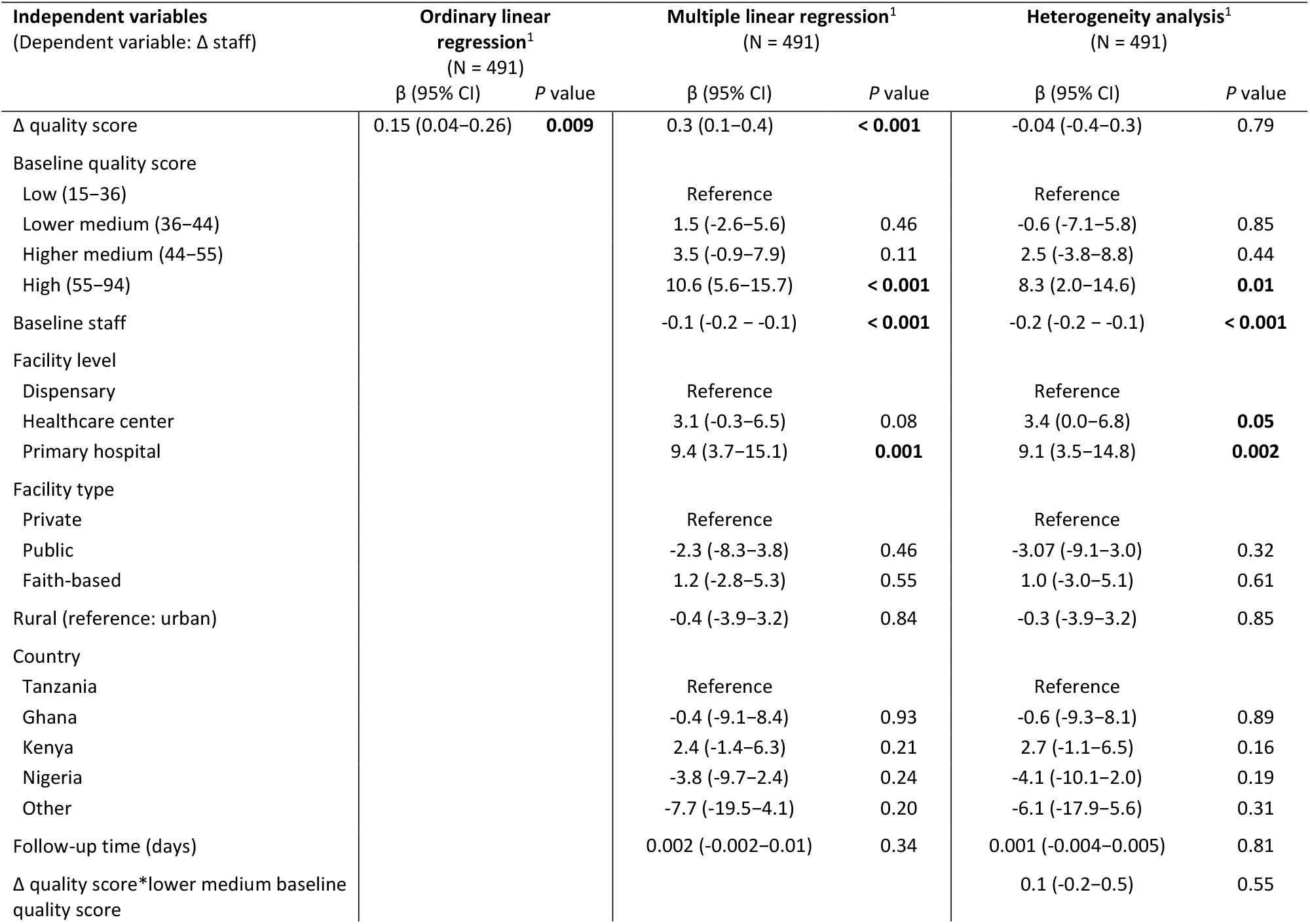
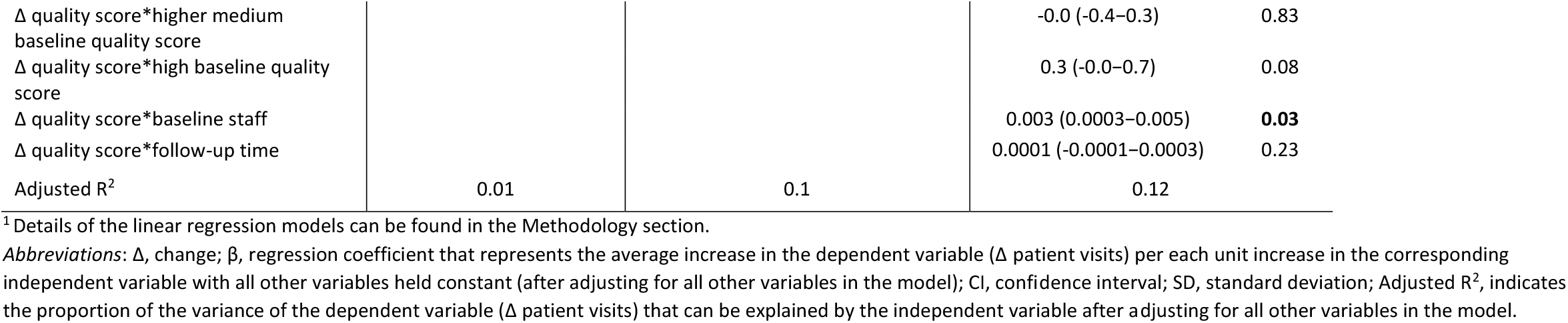
Linear regression models to estimate the effect of the change in quality score on the change in number of staff.

### Improvers versus non-improvers

Table 4 compares the baseline characteristics of the facilities that achieved an improvement of 5 points or higher in the SafeCare quality score at follow-up (n [%], 344 [70.1]), with the facilities that did not achieve this level of improvement (147 [29.9]) classified as the improvers and non-improvers respectively. We found that the improvers had at baseline significantly lower quality scores than the non-improvers (42.6 versus [vs] 51.1, *P* < 0.001) and that their proportion within the low score category was higher (118 [34.3] vs 12 [8.2], *P* < 0.001) (Table 4 Column 1). Regarding facility levels, the proportion of primary hospitals among the improvers was significantly higher (74 [21.5]) versus the non-improvers (20 [13.6], *P* = 0.042). When studying the facilities’ ownership, the proportion of public and faith-based facilities were significantly higher among the improvers versus the non-improvers (Table 4). Finally, the proportion of Tanzanian and Nigerian facilities were significantly lower among the improvers (Table 4).

**Table 4.** Baseline facility characteristics for improvers versus non-improvers.

| Variables | Improvers<sup>1</sup><br>(N = 344) | Non-improvers<sup>2</sup><br>(N = 147) | P value |
| --- | --- | --- | --- |
| # Patient visits, mean (SD) | 1163.78 (1669.6) | 1013.84 (1610.5) | 0.358 |
| # Staff, mean (SD) | 35.56 (48.3) | 27.01 (43.2) | 0.065 |
| Score at baseline, mean (SD) | 42.55 (11.5) | 51.07 (12.5) | **<0.001** |
| Quality score categories, n (%) |  |  |  |
| Low (15–36) | 118 (34.3) | 12 (8.2) | **<0.001** |
| Lower medium (36–44) | 90 (26.2) | 37 (25.2) | 0.819 |
| Higher medium (44–55) | 77 (22.4) | 46 (31.3) | **0.037** |
| High (55–94) | 59 (17.2) | 52 (35.4) | **<0.001** |
| Facility level, n (%) |  |  |  |
| Dispensary | 144 (41.9) | 65 (44.2) | 0.629 |
| Healthcare center | 126 (36.6) | 62 (42.2) | 0.248 |
| Primary hospital | 74 (21.5) | 20 (13.6) | **0.042** |
| Facility type, n (%) |  |  |  |
| Private | 26 (7.6) | 12 (8.2) | 0.819 |
| Public | 185 (53.8) | 101 (68.7) | **0.002** |
| Faith-based | 133 (38.7) | 34 (23.1) | **0.001** |
| Rural, n (%) | 128 (37.2) | 50 (34.0) | 0.501 |
| Country, n (%) |  |  |  |
| Tanzania | 195 (56.7) | 61 (41.5) | **0.002** |
| Ghana | 12 (3.5) | 3 (2.0) | 0.394 |
| Kenya | 109 (31.7) | 60 (40.8) | 0.051 |
| Nigeria | 20 (5.8) | 23 (15.6) | **<0.001** |
| Other | 8 (2.3) | 0 (0) | 0.063 |

<sup>1</sup>Improvers, facilities that had at the follow-up a $\Delta$ quality score $\geq 5$ .
<sup>2</sup>Non-improvers, facilities that had at the follow-up a $\Delta$ quality score $< 5$ .
Abbreviations: $\Delta$ , change; SD, standard deviation.

## Discussion

UHC will produce better health outcomes in LMICs only on the foundations of a high-quality health system (Kruk *et al*. 2018b). In Africa, among the biggest impediments to improve quality of care in small-and medium-size public and private facilities is the limited investment in quality-of-care improvements and regulation from the healthcare authorities. As a result, millions of people die every year in LMICs due to the poor quality of care they receive (Kruk *et al*. 2018c). Evidence indicates that substandard care wastes significant economic and human resources due to for instance duplicated services, ineffective care, and avoidable hospital admissions (WHO 2018a). Hence, good quality health services not only ensure healthier societies but also healthier economies. It is a fallacy that quality of care is a luxury only rich countries can afford, since LMICs, especially the poorest ones, cannot afford the high cost of lack of quality care (WHO 2018a).

To strengthen the investment of private providers in their own facilities, as well as create efficiency in care delivery, private (not-)for-profit facilities need to attract more clients to increase revenues to invest in quality improvement. In turn, higher quality may lead to increased retention of patients, further enhancing revenues. But in a facility with limited resources the question arises whether these investments are worth it. Would small and medium facilities indeed realize an improvement in business performance when investing in quality improvement?

Here, we quantify the (potential) business case for quality improvement in SSA by analysing the quality improvement journey of almost 500 facilities in 7 African countries and their business performance as measured by changes in patients visits and staff between two patients and staff over time matched to changes in quality assessments as measured by SafeCare scores. Our findings point towards an S-shaped association between quality and business performance improvements. Facilities with lower quality scores at baseline, did not materialize a significant change in number of patients visits or staff even if they significantly improved their SafeCare quality score. On the other hand, facilities with a relatively high-quality baseline score (≥ 55), saw a significant increase in the number of patients and staff at 18 months of follow-up. This shows clinics need to reach a certain threshold of quality before further improvements lead to increases in patient utilization, likely fuelled by increased trust and word-of-mouth recommendations of clients, that subsequently translate into a higher demand for services and larger revenues.

These findings pose a dilemma for financing quality improvement within African healthcare systems that should be of concern for policy-makers and other stakeholders. In our cohort of 492 facilities, more than a quarter had a SafeCare score between 15–36 at baseline (low quality score category) (Table 1). When facilities have such low scores, decision-makers might be induced to close them, when at the same time these facilities may be the only provider for many of the surrounding communities. Whereas many of these facilities may have worked hard and significantly improved their score over time, this did not translate yet into an improvement in business performance and a return on investment. A critical infrastructure, and a basic level of human resources are needed to transform quality improvement into good business performance, which is reflected in the higher proportion of primary hospitals among the improvers versus the non-improvers. Therefore, financial support and continued technical assistance may be needed for these facilities to break out this low level of performance. In addition, better contracting, regulation, central supply management, and innovative models will enable them to continue their quality improvement journey, to prevent harming the patients they attend, and to help them succeed as small/medium businesses and keep investing in quality. Improving the quality of care leads to a higher willingness of patients to pay for the healthcare services they receive, which in turns will enhance the capacity to invest of the facilities.

The local private health sector is often underutilized in SSA UHC strategies. This despite its pivotal role in serving half of the population (2008). For instance, the successful public-private collaborations taking place during the COVID-19 pandemic in Africa have led local policy-makers propose it to become the norm in health interventions (WHO 2021). Public-private partnerships have several advantages that include joining complementary economic and human resources, but also to make technology and know-how mutually available and create more efficient referral systems. In order to achieve UHC and high quality of care, it is essential to include the private sector.

To finance quality improvements, digital loans may be an affordable and accessible financing alternative to the traditional loans from banks or government organizations since many facility’s owners do not have the collateral required to apply for bank loans (MCF 2021). Digital loans instead do not require assets and have a flexible and transparent re-payment method. Furthermore, during times of public health emergencies, such as the COVID-29 pandemic, that disrupt the functioning of the financial system, digital loans are able to provide continued financial support to healthcare facilities.

One size does not fit all. Quality improvement programs in LMICs should be customized to the needs of each geographical setting with a clear eye on availability of resources. When available, budgets are limited. Investments on quality improvement could concentrate on those (public and private) healthcare providers that surpass certain thresholds of quality and where senior staff demonstrate positive attitudes towards quality improvement. In addition, smaller facilities have the capacity to attend only to a limited number of patients and might not be able to improve in business performance without investment in scale and scope. Importantly, policy-makers should ensure adequate regulation and effective contracting to certify that key quality areas are targeted and sustainably addressed.

A key limitation of this study is that the facilities included in this analysis were all willing to enrol in the SafeCare program, and therefore they may not be representative of other facilities in similar settings since they have shown a certain interest in quality improvement. Finally, the implementation of quality processes and plans involve a behavioural change. The coaching required to implement the quality improvement plans demand sufficient human resources and may be too expensive (Das & Hammer 2014; Quaife *et al*. 2021; Ugo *et al*. 2016). On the one hand, this may exclude the smallest facilities with least resources. On the other hand, this might imply that learnings from quality improvement assessments are not automatically translated into quality improvements (Luty *et al*. 2022). This is also reflected in the big gaps found in the literature between provider knowledge and practice (Das *et al*. 2015; Powell-Jackson *et al*. 2020). The implementation of such processes can be importantly influenced by the intention to improve the quality of care that for some facilities is driven by altruistic motivation and a sense of duty, while for others such changes may be seen as sound financial investment.

## Conclusions

There is a business case for quality improvement in SSA that can be attained after achieving a certain level of quality, and that can materialize in an increase in revenues from growing patient utilization and an increase in staff numbers. Facilities that have started to implement quality improvements but that are still below a certain threshold, need financial support to accelerate their progress and reach the required quality level beyond which their business performance can further improve. Public-private partnerships, and innovative loan schemes could help these facilities realize their own business case for quality improvement in SSA.

## Data Availability

All data produced in the present study are available upon reasonable request to the authors.

## END MATTER

### Data availability statement

The data sets used and/or analysed during the current study are available from the SafeCare programme at PharmAccess Foundation upon request.

### Funding

This work was supported by the Health Insurance Fund of the Minister of Foreign Affairs of The Netherlands.

#### Acknowledgments

We are grateful to all quality improvement assessors working for SafeCare in Africa, for their indefatigable work in driving quality improvement changes and saving through these changes thousands of lives; to Alex Italiaander for his data management support; to all the facilities enrolled in the SafeCare program and being part of this study.

### Authors contributions

AdG, NS, WJ, TRdW conceived and designed the study; DB, performed the statistical analysis; GPGP, wrote the first draft of the manuscript; JD and PR provided technical assistance regarding SafeCare data used for this analysis; all authors have reviewed and approved the final version of the manuscript.

### Ethical approval

Ethical approval for this type of study is not required by our institute.

## References

2008. The World Bank - The business of health in Africa : partnering with the private sector to improve people’s lives. https://documents.worldbank.org/en/publication/documents-reports/documentdetail/878891468002994639/the-business-of-health-in-africa-partnering-with-the-private-sector-to-improve-peoples-lives, accessed 10 May 2021. 2017. The World Bank - Population Data

2018. The World Bank - Current health expenditure per capita (current US$). https://data.worldbank.org/indicator/SH.XPD.CHEX.PC.CD, accessed 12 July 2021.

Anyangwe SC, Mtonga C. 2007. Inequities in the global health workforce: the greatest impediment to health in sub-Saharan Africa. Int J Environ Res Public Health, 4: 93–100.

Das J, Hammer J. 2014. Quality of Primary Care in Low-Income Countries: Facts and Economics. Annual Review of Economics, 6: 525–553.

Das J, Kwan A, Daniels B, et al. 2015. Use of standardised patients to assess quality of tuberculosis care: a pilot, cross-sectional study. Lancet Infect Dis, 15: 1305–13.

Johnson MC, Schellekens O, Stewart J, et al. 2016. SafeCare: An Innovative Approach for Improving Quality Through Standards, Benchmarking, and Improvement in Low- and Middle-Income Countries. Jt Comm J Qual Patient Saf, 42: 350–71.

Jowett M Pbm, Flores G, Cylus J. 2016. Spending targets for health: no magic number - WHO and Health Systems Governance & Financing http://apps.who.int/iris/bitstream/handle/10665/250048/WHO-HIS-HGF-HFWorkingPaper-16.1-eng.pdf?sequence=1, accessed 12 July 2021.

Kruk ME, Gage AD, Arsenault C, et al. 2018a. High-quality health systems in the Sustainable Development Goals era: time for a revolution. Lancet Glob Health, 6: e1196–e1252.

Kruk ME, Gage AD, Arsenault C, et al. 2018b. High-quality health systems in the Sustainable Development Goals era: time for a revolution. Lancet Glob Health, 6: e1196–e1252.

Kruk ME, Gage AD, Joseph NT, et al. 2018c. Mortality due to low-quality health systems in the universal health coverage era: a systematic analysis of amenable deaths in 137 countries. Lancet, 392: 2203–2212.

Luty JT, Oldham H, Smeraglio A, et al. 2022. Simulating for Quality: A Centralized Quality Improvement and Patient Safety Simulation Curriculum for Residents and Fellows. Acad Med, 97: 529–535.

MCF. 2021. Healthcare businesses in Africa to benefit from a digital finance fund. https://www.medicalcreditfund.org/update/healthcare-businesses-in-africa-to-benefit-from-a-digital-finance-fund/, accessed April 7 2022.

Onno P. Schellekens MEL, Joep M.A. Lange, Jacques van der Gaag. 2007. A new paradigm for increased access to healthcare in Africa. https://www.pharmaccess.org/wp-content/uploads/2020/04/A-new-paradigm-for-increased-access-to-healthcare-in-Africa_OS.pdf, accessed May 11 2021.

PharmAccess. 2022. Medical Credit Fund. https://www.medicalcreditfund.org/, accessed February 1 2022.

Powell-Jackson T, King JJC, Makungu C, et al. 2020. Infection prevention and control compliance in Tanzanian outpatient facilities: a cross-sectional study with implications for the control of COVID-19. Lancet Glob Health, 8: e780–e789.

Quaife M, Estafinos AS, Keraga DW, et al. 2021. Changes in health worker knowledge and motivation in the context of a quality improvement programme in Ethiopia. Health Policy Plan, 36: 1508–1520.

Roser M, Ritchie H. 2017. Burden of Disease. Published by: Our World in Data. https://ourworldindata.org/burden-of-disease#the-global-distribution-of-the-disease-burden, accessed 28 April 2021.

Spieker N. 2020. The Business Case for Quality in Health Care. In: Marquez LR (ed). Improving Health Care in Low- and Middle-Income Countries: A Case Book. Springer.

Ugo O, Ezinne EA, Modupe O, et al. 2016. Improving Quality of Care in Primary Health-Care Facilities in Rural Nigeria: Successes and Challenges. Health Serv Res Manag Epidemiol, 3.

WHO. 2018a. Delivering quality health services: a global imperative for universal health coverage. https://apps.who.int/iris/handle/10665/272465, accessed May 10 2021.

WHO. 2018b. State of health in the WHO African Region. https://www.afro.who.int/publications/state-health-who-african-region, accessed May 10 2021.

WHO. 2021. WHO’s results in Africa between July 2020 – June 2021. Report of the Regional Director. chrome-extension://efaidnbmnnnibpcajpcglclefindmkaj/viewer.html?pdf url=https%3A%2F%2Fwww.afro.who.int%2Fsites%2Fdefault%2Ffiles%2F2021-08%2F016_WHO-AFRO_RD-Report-20-21_Ex-Summary_EN_0.pdf&clen=2655585&chunk=true, accessed February 2 2022.

World Health Organization. 2019. Integrating Health in Africa: The Role of the Private Sector. https://www.afro.who.int/regional-director/speeches-messages/integrating-health-africa-role-private-sector, accessed 10 May 2021.

