## Supplementary Table 2 for "The Business of Quality: Evaluating the Impact of Healthcare Quality Improvements on Staff Numbers and Patient Utilization in Sub-Saharan Africa"

**Supplementary Table 2**. Linear regression models to estimate the effect of the change in business performance quality scores on the change in number of staff.

| **Independent variables**  (Dependent variable: Δ staff) | **Ordinary linear regression**^1^  (N = 491) | | | **Multiple linear regression**^1^  (N = 491) | | | **Heterogeneity analysis**^1^ (N = 491) | |
| --- | --- | --- | --- | --- | --- | --- | --- | --- |
|  | β (95% CI) | *P* value | | β (95% CI) | | *P* value | β (95% CI) | *P* value |
| Δ quality score | 0.16 (0.06−0.25) | | **0.001** | 0.2 (0.1−0.3) | **0.001** | | 0.1 (0.0−0.3) | 0.06 |
| Baseline quality score |  |  | |  | |  |  |  |
| Low (1.0−5.0) |  |  | | Reference | |  | Reference |  |
| Lower medium (5.5−11.0) |  |  | | 0.6 (-4.0−5.1) | | 0.81 | 1.4 (-4.2−6.9) | 0.62 |
| Higher medium (11.3−27.3) |  |  | | 1.3 (-3.3−5.9) | | 0.59 | 1.5 (-4.4−7.4) | 0.61 |
| High (27.5−73.0) |  |  | | 5.8 (0.2−11.3) | | **0.04** | 8.9 (2.0−15.8) | **0.01** |
| Baseline staff |  |  | | -0.11 (-0.16 − -0.7) | | **< 0.001** | -0.16 (-0.2 − -0.1) | **< 0.001** |
| Facility level |  |  | |  | |  |  |  |
| Dispensary |  |  | | Reference | |  | Reference |  |
| Healthcare center |  |  | | 2.9 (-0.6−6.5) | | 0.11 | 2.7 (-0.9−6.2) | 0.14 |
| Primary hospital |  |  | | 8.7 (2.7−14.7) | | **0.005** | 8.0 (2.0−14.0) | **0.01** |
| Facility type |  |  | |  | |  |  |  |
| Private |  |  | | Reference | |  | Reference |  |
| Public |  |  | | -3.1 (-9.4−3.2) | | 0.34 | -3.6 (-9.9−2.8) | 0.28 |
| Faith-based |  |  | | 1.0 (-3.1−5.2) | | 0.62 | 0.6 (-3.5−4.8) | 0.77 |
| Rural (reference: urban) |  |  | | -0.1 (-3.8−3.5) | | 0.94 | 0.1 (-3.5−3.8) | 0.95 |
| Country |  |  | |  | |  |  |  |
| Tanzania |  |  | | Reference | |  | Reference |  |
| Ghana |  |  | | 0.9 (-8.1−9.9) | | 0.84 | 1.0 (-8.0−10.0) | 0.83 |
| Kenya |  |  | | 2.1 (-1.9−6.1) | | 0.31 | 2.6 (-1.5−6.6) | 0.21 |
| Nigeria |  |  | | -2.4 (-8.8−4.1) | | 0.48 | -1.8 (-8.3−4.6) | 0.58 |
| Other |  |  | | -8.4 (-20.7−3.8) | | 0.18 | -6.7 (-19.1−5.7) | 0.29 |
| Follow-up time (days) |  |  | | 0.003 (-0.001−0.01) | | 0.17 | 0.002 (-0.003−0.006) | 0.47 |
| Δ quality score*lower medium baseline quality score |  |  | |  | |  | -0.1 (-0.4−0.2) | 0.57 |
| Δ quality score*higher medium baseline quality score |  |  | |  | |  | -0.1 (-0.4−0.3) | 0.57 |
| Δ quality score*high baseline quality score |  |  | |  | |  | -0.3 (-0.7−0.1) | 0.11 |
| Δ quality score*baseline staff |  |  | |  | |  | 0.004 (0.001−0.006) | **0.004** |
| Δ quality score*follow-up time |  |  | |  | |  | 0.0001 (-0.0001−0.0003) | 0.43 |
| Adjusted R^2^ | 0.01 | | | 0.09 | | | 0.08 | |

^1^ Details of the linear regression models can be found in the Methodology section. For the models described in this table, only the quality scores of the criteria related to business performance were included in this analysis, which comprises 38 out of a total of 753 SafeCare criteria.

*Abbreviations*: Adjusted R2, indicates the proportion of the variance of the dependent variable (Δ patient visits) that can be explained by the independent variable after adjusting for all other variables in the model; Δ, change; β, regression coefficient that represents the average increase in the dependent variable (Δ patient visits) per each unit increase in the corresponding independent variable with all other variables held constant (after adjusting for all other variables in the model); CI, confidence interval; SD, standard deviation.
